# Feasibility of individualized home exercise programs for patients with head and neck cancer – study protocol and first results of a multicentre single-arm intervention trial (OSHO #94)

**DOI:** 10.1101/2024.03.17.24304427

**Authors:** Sabine Felser, Julia Rogahn, Änne Glass, Lars Arne Bonke, Daniel F. Strüder, Jana Stolle, Susann Schulze, Markus Blaurock, Ursula Kriesen, Christian Junghanss, Christina Grosse-Thie

## Abstract

**Introduction:** Patients with head and neck cancer (PwHNC) benefit from targeted exercise interventions: symptom relief, compensation for dysfunction, improvement in quality of life (QoL). Data on acceptance physical interventions in PwHNC are rare. The ‘OSHO #94’ trial investigates the short- and medium-term effects of individualized home exercise in PwHNC on QoL, physical activity and functionality. The study includes a feasibility phase in order to evaluate the acceptance (Phase A), followed by a consecutive QoL endpoint Phase B. Here we present the study protocol as well as the feasibility results.

**Methods and analysis:** This prospective, multicentre, single-arm intervention study includes PwHNC ≥18 years of age in aftercare or palliative care with stable remission under immunotherapy. The study opened in January 01, 2021, with estimated completion by December 31, 2024. The PwHNC receive an individualized home exercise program consisting of mobilization, coordination, strengthening and stretching exercises. This should be carried out at least three times a week over 12 weeks for 15 to 30 minutes, supplemented by aerobic training two to three times a week for 30 minutes (intervention). Once weekly telephone calls with a physiotherapist are performed. Subsequently, there is a 12-week follow-up (FU) without exercise specifications/contact. Outcomes are measured before and after the intervention and following the FU. Primary outcome of the feasibility phase (Phase A, n = 25) was the determination of the dropout rate during the intervention with a termination cut off if more than 30% PwHNC withdrew premature. The primary outcome of phases A + B (N = 53) are the change in global QoL score from pre- to post-intervention (EORTC QLQ-C30). Secondary outcomes include clinical and patient-reported measures, training details as well as functional diagnostic data (e.g. level of physical activity, training frequency, flexibility, fall risk and aerobic performance).

**Results:** 25 PwHNC were enrolled onto the feasibility cohort. Only16% (4/25 patients) did not complete the study. Therefore, individualized home exercise programs in PwHNC seem feasible recruitment of PwHNC for phase B continued. The dropout rate was adjusted from 30% (N = 60) to 20% (N = 53, calculated sample size n = 42 PwHNC and 20% (n = 11) to dropout).

**Ethics and dissemination:** The study protocol was approved by the Ethics Committee of the University of Rostock, University of Halle-Wittenberg and University of Greifswald. The findings will be disseminated in peer-reviewed journals and academic conferences.

**Trial registration** German Registry of Clinical Trials DRKS00023883.

## Introduction

Head and neck cancer (HNC) comprises various types of cancer that occur in the head and neck area, including malignant neoplasms of the oral cavity, pharynx, larynx, nose and paranasal sinuses. Many patients with HNC (PwHNC) have clinically relevant functional deficits, often in the areas of food-intake, breathing, speech, pain, mood and neck and shoulder mobility, due to the location of the tumours and intensive local therapy procedures (1–4), e.g. operation and/or radiotherapy. In addition, there may be visible disfigurements in the face and neck area, weight loss and sarcopenia (5), body image disturbance (6) or other symptoms such as fatigue. These acute and sometimes chronic disease- and therapy-related functional deficits and side effects often impair the health-related quality of life (QoL) of those affected (5–9). Recent advances in the diagnosis and treatment of PwHNC have significantly improved the survival of PwHNC (10). As a result, there are more long-term survivors (11), and, especially under new approaches such as immunotherapy (ICT-immune-checkpoint therapy), a small group of PwHNC can reach stable remissions even in a primary incurable situation (12). Therefore, other dimensions of the treatment outcome, such as physical status and functional abilities, psychological status and wellbeing, are becoming increasingly important (13). Consequently, the treatment of functional deficits and the improvement of QoL is an essential task in the context of interdisciplinary rehabilitation of PwHNC (1, 2).

The most commonly used supportive interventions in HNC survivors to date have focused on monitoring/treatment of physical effects (14). Physical exercise is a feasible, safe and promising approach to improve QoL in HNC survivors. In particular, 12-week training programs with aerobic activity (walking) or progressive resistance training for the whole body showed great benefit for improving QoL perception in HNC survivors (15). In addition, reviews and meta-analyses indicate that PwHNC benefit from exercise interventions during and after medical therapy in terms of physical functionality (muscle strength, cardiorespiratory fitness, flexibility), (shoulder) pain reduction and fatigue relief. The effects described with regard to body composition are heterogeneous (16–19). Consequently, the American Head and Neck Society’s 2022 statement on exercise therapy calls for, among other things, 1. early screening of PwHNC for rehabilitation needs using, 2. objective assessments, 3. referral of PwHNC to qualified therapists, and 4. motivating and encouraging PwHNC to engage in regular physical activity (20). The fact that particular attention should be paid to the latter point is illustrated by study results from Taiwan and Sweden, which show that PwHNC are insufficiently physically active (21, 22). According to the results of Fang et al. (21), only 17% of 108 HNC survivors met the WHO criteria for physical activity. Those reported less fatigue and better QoL compared to PwHNC who did not meet the criteria. Regardless, the PwHNC had poorer overall physical fitness compared to results from normative data of subjects from Taiwanese work fitness measurements. Reviews of the barriers revealed that in addition to physical problems, time pressure, lack of motivation, and lack of knowledge are the main reasons for insufficient physical activity in PwHNC (23, 24). The results also revealed that the exercise preferences of the PwHNC differ in terms of type, location, company, intensity, frequency and supervision. It is known from surveys in England, the US Midwest and Canada that PwHNC prefer (un)supervised exercise programs, alone or with family members, at moderate intensity, at home or outdoors at different times (25–27). To summarize the current findings, it can be concluded that individual exercise programs that can be carried out flexibly at home and/or outdoors are an optimal approach to motivate PwHNC to be more physically active. However, this approach has not yet been sufficiently investigated. In addition, there is little knowledge about the sustainability of (home) training interventions in PwHNC.

As a result, our goal is to develop an exercise program that can be carried out 100% independently and flexibly by PwHNC at home or outdoors. This exercise program should be designed in a way that it can be flexibly adapted to the individual needs of PwHNC. The short and medium-term effects on QoL, the level of physical activity, physical functionality and body composition will be investigated. Here we present the study protocol and the results of the feasibility study.

## Methods and Analysis

### Study design and, primary and secondary outcomes

OSHO #94 is a prospective, multicentre, single-arm, intervention trial conducted by the Department of Haematology, Oncology and Palliative Care of Rostock University Medical Centre (clinic III, UMR, Germany) in adult PwHNC.

The study consists of two phases, A and B. Phase A examines the feasibility of the study design including the estimation of the dropout rate during the intervention. Further phase A outcomes are compliance with the training recommendations, the number of adverse events associated with the training, satisfaction with the training information, use of the exercise materials and the influence of weekly telephone calls during the intervention on the motivation to train. Once phase A has been successfully completed, which involves a maximum dropout rate of 30%, the study was supposed to automatically enter Phase B. Otherwise, it was planned to modify the study design.

In Phase B, the short- and medium-term effects of the intervention on QoL will be investigated. We hypothesize that the primary outcome ‘global QoL score’ will increase significantly after a 12-week guided training intervention in the home-based setting. Secondary outcomes are the level of physical activity, physical function and body composition.

### Recruitment, and inclusion and exclusion criteria

Recruitment started on January 1, 2021 at the UMR in cooperation with the Department of Otorhinolaryngology, Head and Neck Surgery ‘Otto Koerner’. The Krukenberg Cancer Centre Halle of the University Hospital Halle has been recruiting since September 20, 2022 and the Department of Otorhinolaryngology, Head and Neck Surgery at the University Medicine Greifswald since March 1, 2023. The study team is supported in the application/recruitment process by the self-help network Kopf-Hals-M.U.N.D.-Krebs e. V. Recruitment should be completed on December 31, 2024.

Patients must meet all of the following inclusion criteria: age ≥18 years, a final coded diagnosis according to ICD: C00-C14, C30-C32 (HNC) in aftercare (after antineoplastic therapy or after completion of rehabilitation, if planned) or with stable remission under immunotherapy and medical clearance of the treating physician, able to walk.

The exclusion criteria are inadequate knowledge of the German language, consent not given, clinically relevant heart failure (NYHA III and IV), myocardial infarction within the last 4 weeks, unstable angina pectoris, higher-grade valvular vitia, uncontrolled cardiac arrhythmias, chronic obstructive pulmonary disease (GOLD III and IV), peripheral arterial occlusive disease (≥Stage III according to Fontaine), diseases that could seriously impair cognitive performance (e.g. dementia, stroke, Wernicke-Korsakoff syndrome), known alcohol dependency and score <24 points on the Mini-Mental State Examination (MMSE).

### Preliminary work: conception of a home exercise program for PwHNC

In an initial study conducted in 2018/19 at UMR’s Clinic III, a training program with exercises for PwHNC suitable for independent training at home was developed. Afterwards, this training program was evaluated by means of a 12-week training intervention in a group setting with regard to feasibility (dropout, adherence) and the effects on physical functionality and QoL in a pilot cohort (28). The low dropout rate (17%), the good adherence of the participants (83%) and the positive effects in terms of physical functionality and QoL provided an excellent basis for transferring the training program to home exercise. In preparation for this, an “‘Exercise manual for patients with mouth, jaw, face and throat tumours’” was drafted containing a total of 90 mobilization, coordination, strengthening and stretching exercises for training at home (29). Video clips for these 90 exercises were created in line with the book and supplemented with four exercise videos with the following content: (1.) Functional gymnastics & balance, time approx. 20 min, (2.) Coordination training, time approx. 12 min, (3.) Mobilization, coordination and strength training with exercise ball, time approx. 22 min, (4.) Training of the shoulder and neck region, time approx. 30 min.

### Study procedure

The study procedure is shown schematically in Fig 2. Screening of outpatients who come to the consultation is carried out locally at the recruiting study centres. Potential study patients are approached directly by the study team and informed about the study. PwHNC who have become aware of the study through the self-help network Kopf-Hals-M.U.N.D.-Krebs e. V. or through flyers can also contact the study centres. If the patient meets the inclusion criteria without any exclusion criteria, the patient is informed by a physician or sports scientist/physiotherapist. Following written consent by the patient, the pre-examination is administered. Based on the results of this examination and the objectives of the PwHNC, the therapists create an individual training plan. Depending on the patientś place of residence, time and state of health, they will be introduced to the training program following the pre-examination or at a separate appointment within the next two weeks. Study patients receive the above-mentioned exercise manual for training at home. In this manual, the exercises recommended by the therapists are marked with a green sticky dot, and contraindicated exercises are marked with a red sticky dot. If the patients have the appropriate technology (e.g. PC) and wish to do so, the therapists put the recommended exercises together in a video clip. These are stored together with selected exercise programs, the four mentioned above, on a USB stick and given to the patients. Patients also receive a free elastic band for strengthening exercises and an inflatable exercise ball (Ø 22 cm) for coordination and strengthening exercises at home. The 12-week individual home exercise program starts immediately after admission. During the 12-week intervention, the therapists contact the patients by telephone once a week. In a telephone protocol, they record the training completed (type, frequency and time), adverse events associated with the training, the patient’s motivation to train and the current well-being of the PwHNC. Any questions the patients may have about the training are answered and, if necessary, further training tips are given. Patients who end the intervention prematurely are asked to state the reason for doing so. In addition to the telephone protocol, patients are asked to keep a training diary for the whole intervention period. Following the 12-week home exercise program, the post-examination takes place. After the post-examination, the patients are left to their own resources on the assumption that they can carry out the exercise program or other physical activities on their own without further instruction (= Follow up, FU). After a further 12 weeks, an FU-examination is carried out in order to be able to make statements about the medium-term effects of the home exercise intervention.

**Figure 1.**
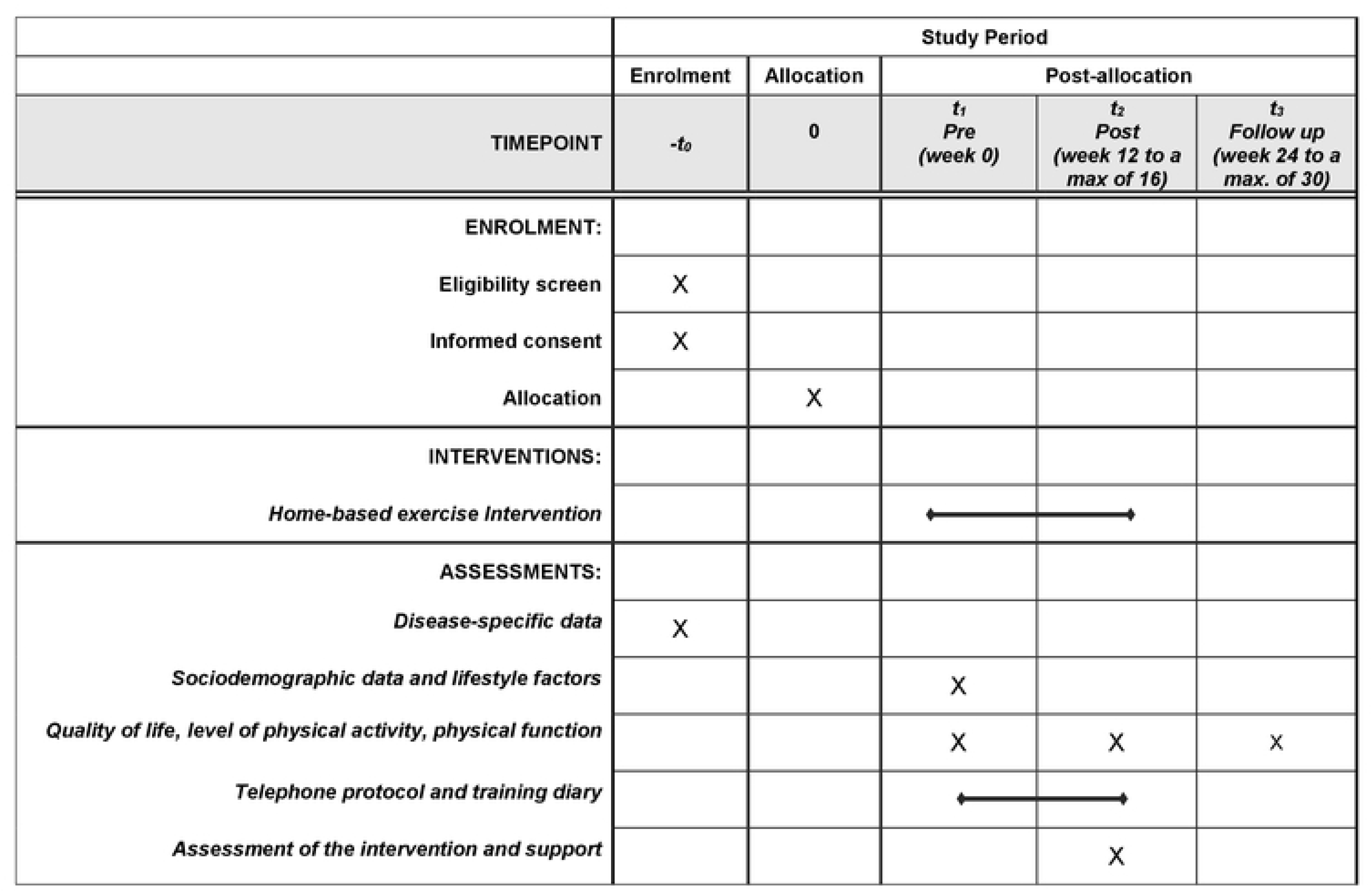
SPIRIT schedule of enrolment, interventions, and assessments

**Fig 2.**
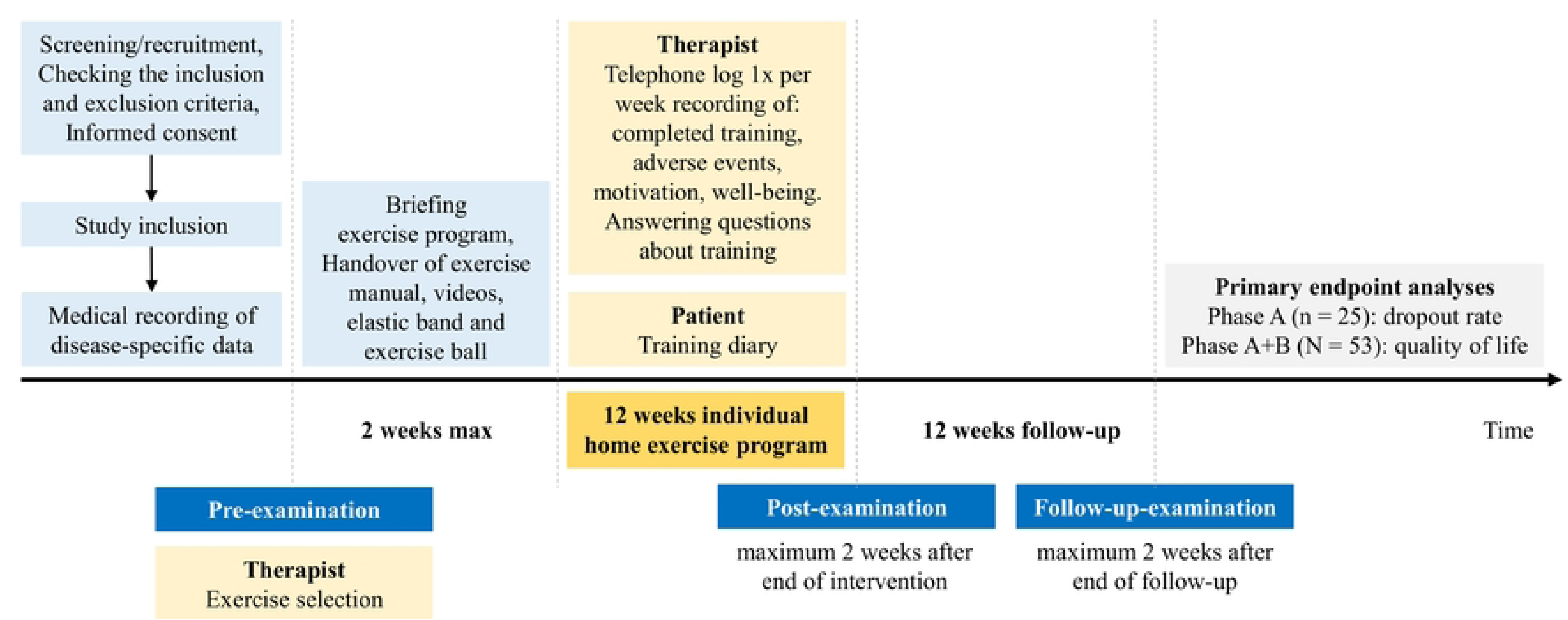
Study procedure OSHO #94

### Exercise recommendations

The training recommendations include completion of the individual exercise program or alternatively one of the four exercise videos on at least 3 days per week for 15–30 minutes. The individual training programs consist of a selection of 15–25 exercises. Depending on the participant’s current fitness level and needs, it includes a different number of mobilization, coordination, strengthening and stretching exercises. The recommended duration of each exercise varies between 30 and 60 seconds and from 1 to 3 repetitions/sets. In addition, endurance training, e. g. (Nordic) walking, cycling, swimming, dancing or similar, is recommended at a frequency of 2 to 3 times a week for 30 minutes each time. The training intensity is controlled using the 15-point BORG rating of perceived exertion scale (30), and the recommended range is between 11 (fairly easy) and 15 (hard).

### Outcome measures

The data collection schedule is shown in Table 1.

**Table 1.**
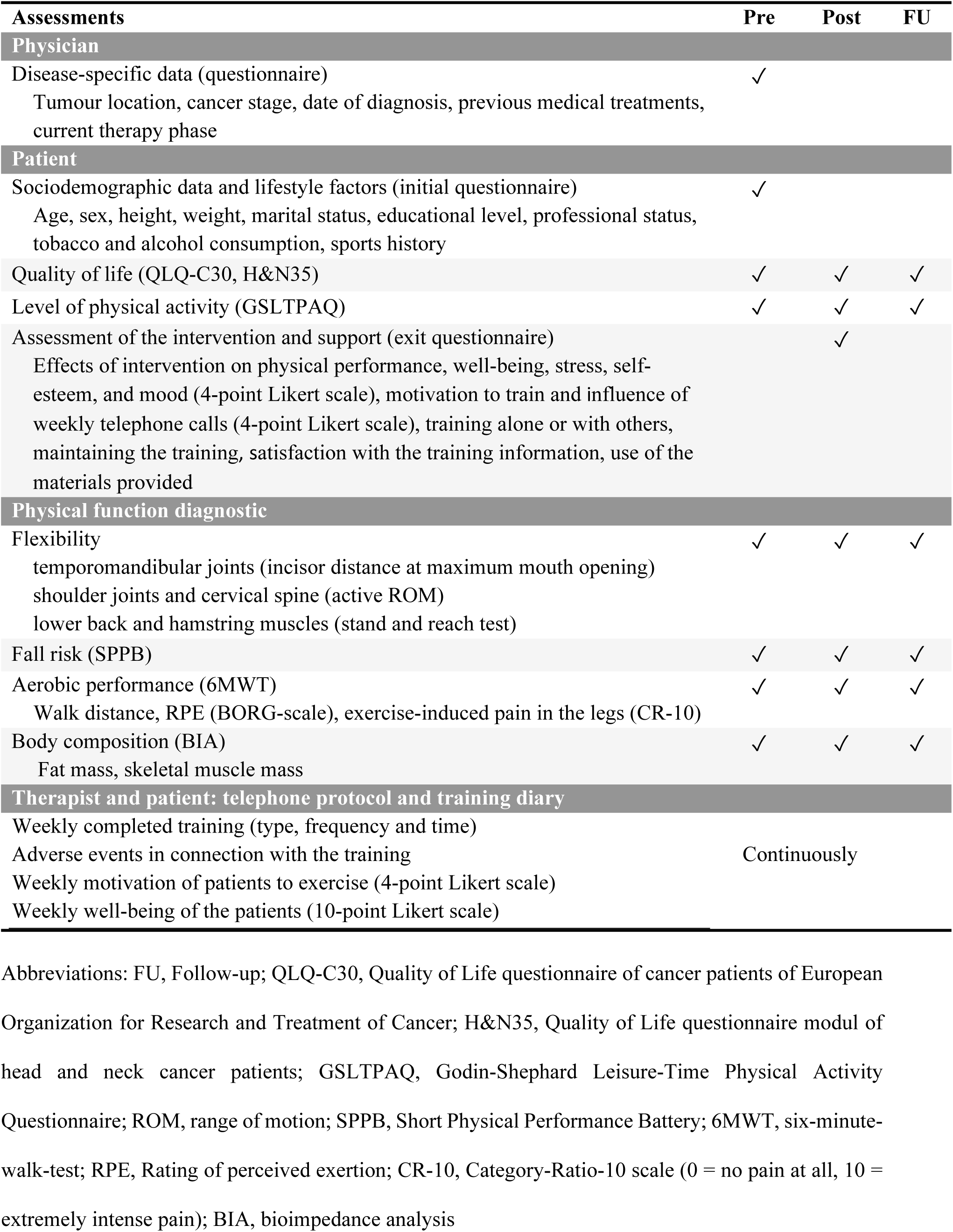
Data collection schedule

*Disease-specific data* (tumour location, cancer stage, date of diagnosis, previous medical treatments and current therapy phase) on the study patients are recorded once by the treating/including physicians at the time of study inclusion.

At the three examination appointments – pre (week 0), post (week 12 to a maximum of 16) and FU (week 24 to a maximum of 30) – patient-reported outcomes (PROs) are recorded, and a physical function diagnostic is carried out. The assessment consists of the following measures:

Patient-reported:

a. *Sociodemographic data* (age, sex, height, weight, marital status, educational level, professional status), and *lifestyle factors* (tobacco and alcohol consumption, sports history) are recorded using an initial questionnaire. The body mass index (BMI) is calculated (body weight [kg]/height [m^2^]).
b. *QoL* is measured using two established questionnaires: (1) EORTC QLQ-C30 version 3.0 questionnaire and (2) the QLQ-HN35 head and neck–specific questionnaire (31). The EORTC QLQ-C30 is a 30-item cancer-specific questionnaire that has a global QoL scale, 5 functional scales, 3 symptom scales and 6 single items. EORTC QLQ-HN35 is a 35-item module. It contains 7 symptom scales and 6 symptom items. Each scale results in an average score of 0 to 100. A high value on the scale ‘global QoL’ and on the functional scales means a high degree of subjectively perceived health and a high assessment of the QoL or a high degree of performance and function. A high value in the symptom scales correlates with a high degree of complaints and symptoms (32–34). No threshold values were set in advance to assess the clinically important difference. The primary outcome is determined by comparing the global QoL score pre- and post-intervention.
c. *Level of physical activity* is assessed using the Godin-Shephard Leisure-Time Physical Activity Questionnaire (GSLTPAQ) (35, 36). The GSLTPAQ is a 4-item self-administered questionnaire. The first three questions seek information on the number of times one engages in mild, moderate and strenuous physical activity bouts of at least 15 min duration in a typical week (37). Scores derived from the GSLTPAQ include total weekly leisure-time physical activity, called a Leisure Score Index (LSI), in which number of bouts at each intensity is multiplied by 3, 5 and 9 metabolic equivalents and summed. LSI scores can be used for ranking individuals from the lowest to highest physical activity levels (38).
d. *Assessment of the intervention and support* is carried out in an exit questionnaire. First, the study participants assess the effects of the intervention on physical performance, well-being, stress, self-esteem, and mood on a 4-point Likert scale. Second, how difficult it was for the PwHNC to motivate themselves to train and what influence the weekly physiotherapy phone calls had on their motivation to train were also recorded on a 4-point Likert scale. Third, it is recorded whether the PwHNC carried out their training alone or with others (e. g. family members). Fourth, the PwHNC are asked whether they will maintain the training. Fifth, the PwHNC were asked whether the information they received about the training was sufficient and, if not, what additional information they would have liked. Sixth, the PwHNC should indicate whether they used the exercise manual, the videos and the small equipment provided for their training.

Physical function diagnostic:

Physical function assessments are carried out a maximum of two weeks before the intervention and a maximum of two weeks after the intervention and the FU. These are performed in the same order. The aim is for all tests to be carried out at one location by the same investigator. The assessment consists of the following measures:

*The flexibility of the temporomandibular joints* is measured using the distance between the incisors (cm) determined with a ruler at maximum mouth opening.
*The flexibility of the shoulder joints and the cervical spine* is measured by the active range of motion (ROM) in the sagittal, frontal, and transversal planes using a manual goniometer. The measurements are carried out starting from the maximum ROM away from the body to the end position close to the body (°).
*The flexibility of the lower back and hamstring muscles* is assessed with the stand and reach test.

Subjects stand with closed/stretched legs and hold one (39) hand covering the other. The trunk is slowly flexed, and the distance between the hands and the ground (which can be hold for 2 s) is measured (cm) (39).

*Fall risk* is evaluated by using the short physical performance battery (SPPB). The SPPB has been shown to have predictive value for the assessment of mortality risk, nursing home admission and disability (40). The SPPB is a group of measures that combines the results of balance tests, gait speed and repeated chair stands. The scores range from 0 (worst performance) to 12 (best performance).
*Aerobic performance* is assessed using the 6-minute walk test (6MWT) (41). The primary measure is the walk distance (m) achieved within 6 minutes. The test is carried out on a 40-m long straight track (a mark is placed every 10 m) on solid ground. After the 6MWT, the participants are asked to rate their perceived exertion (RPE) using the 15-point Borg scale (6 = really, really easy, 20 = maximum effort) (30) and their exercise-induced pain in the leg muscles using a Category-Ratio (CR)-10 scale (0 = no pain at all, 10 = extremely intense pain) (42).

Following completion of the intervention, the telephone protocols and training diaries are evaluated with regard to the following parameters:

*Compliance with regard to training recommendations*: the documented training sessions are evaluated with regard to the average weekly training frequency and time, separately for individual exercise programs and endurance training.
*Adverse events* in connection with the intervention that are mentioned by the participants are recorded.
*The patients’ motivation to train* is recorded over the 12-week intervention using a 4-point Likert scale (1 = not at all, 4 = very much).
*Weekly well-being* is documented and analysed on a 10-point Likert scale (0 = very poor, 10 = very good).

If possible, the reason for dropping out is asked in the event of early termination of studies.

### Statistical considerations

#### Estimation of sample size

The sample size calculation for the trial (phase B) is based on the results of Felser et al. (28): global QoL pre: 50.1 ± 16.4; post: 58.3 ± 16.2; r = 0.618 resulting in an effect size of 0.5755, which is detectable on a confidence level of 1 − α = .95 (2-sided) with n = 42 patients (nQuery® Advisor 7.0 Statistical Solutions Ltd., Boston, MA, USA). Assuming a 20% dropout rate based on our experience, a total of N = 53 patients is required to be included in the study.

#### Statistical analysis

Quantitative variables (i.e. QoL) are presented as mean ± standard deviation or as median (Q1, Q3), ranging from minimum to maximum (min to max); qualitative ones as relative frequency of their occurrence % and absolute (n). Missing data are indicated but not included in the calculation of percentage. The normal distribution of the data is checked using the Shapiro-Wilk test.

*Data analysis phase A:* The estimated dropout rate is given with the respective 95% confidence interval (95%CI) for feasibility. Further outcomes are patient characteristics, the completed training volume, adverse events associated with the training, satisfaction with the training information, use of the exercise materials and the influence of the weekly telephone calls during the intervention phase on motivation. *Data analysis phase B:* QoL, physical activity level, physical functionality and body composition will be measured at three time points (pre, post, FU). Subgroup analyses are planned according to the patient characteristics, including men vs. women, sports beginners vs. PwHNC with sports experience, low vs. high physical performance/ physical activity level/ QoL and centres. Differences in means between time points and groups are analysed by using repeated measures analysis of variance. Correlation analyses are carried out to examine the strength and direction of relationships between quantitative variables. Regression analysis will be used to quantify the influence of predictors on QoL or physical function (flexibility, fall risk, aerobic performance) in a multiple approach.

A *p*-value of <0.05 is considered significant. All data will be analysed using IBM^®^ SPSS^®^.

### Patient involvement

UMR’s clinic III works closely with the local self-help group and the German self-help network Kopf-Hals-M.U.N.D.-Krebs e. V. to educate PwHNC and their families about physical activity and sports. The local self-help group contributed from the beginning to the design and implementation of the OSHO #94 study by helping to create the exercise manual (29) and videos. Both the local self-help group and the self-help network Kopf-Hals-M.U.N.D.-Krebs e. V. inform PwHNC about OSHO #94 via social media and information events. On patient days, study participants report on their experiences with the exercise program, and interim results are presented and discussed together.

### Ethics and Dissemination

The study protocol (version 1 from September 30, 2020) was approved by the Ethic Committee of the University of Rostock (A2020-0274, October 26, 2020). Version 2 from June, 13, 2022 and version 3 from July, 19, 2022 was approved by the Ethics Committee of the University of Halle-Wittenberg (2022-067, June 30, 2022) and University of Greifswald (BB 117/22, September 13, 2022). Three protocol modifications have been made and approved so far. These included the extension of the recruitment period twice (amendments from December 2021, and March 2024), the inclusion of PwHNC under palliative care (amendment from July 2022), and the change of deputy study leader (amendment from March 2024). The study was carried out in accordance with the Declaration of Helsinki and is registered at the German Clinical Trials Register (DRKS00023883). All participants will have to sign and date an informed consent form. The findings will be disseminated in peer-reviewed journals and academic conferences.

## Results of feasibility (phase A)

Between January 2021 and February 2023, 25 PwHNC were included in the study. The proportion of men was 52% (n = 13). The median age was 66 (61, 72), ranging from 20 to 85 years. A higher education degree (>10 years) was held by 60% (n = 15) of the participants, 56% (n = 14) were non-smokers and 72% (n = 18) stated that they had been active in sports before the disease. The most common tumour location was the oropharynx (32%, n = 8), followed by the oral cavity (28%, n = 7). Participants were first diagnosed between 2006 and 2022. At the time of study participation, 92% (n = 23) were in complete remission. Further details on the socio-demographic, lifestyle and clinical data of the study participants can be found in Table 2.

**Table 2.**
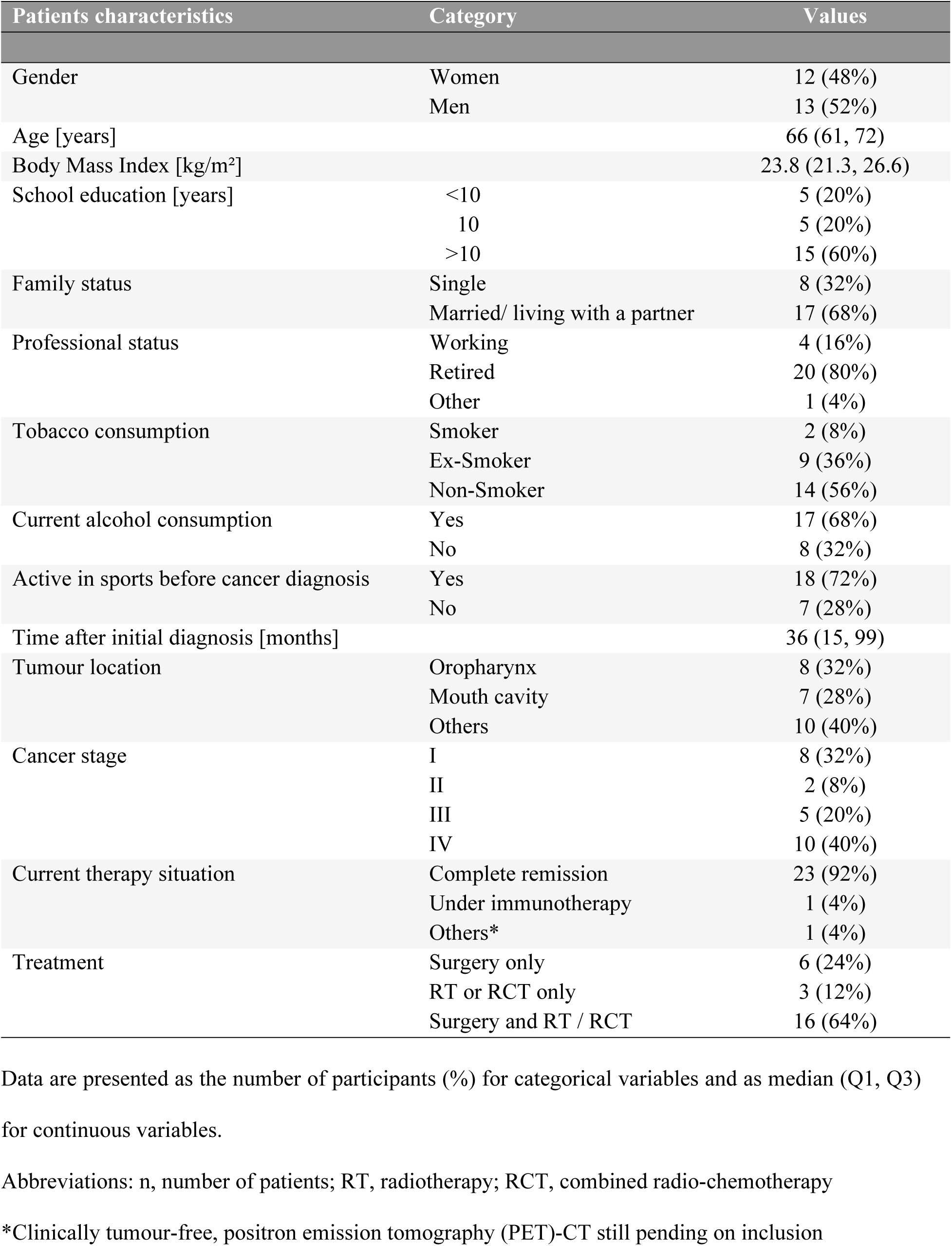
Sociodemographic and clinical data (n = 25)

### Dropout rate

A total of 22 out of 25 participants completed the intervention. Three participants (12%) discontinued the intervention prematurely for health reasons (operations, psychological stress due to unclear findings). Data from 21 participants were analysed due to the fact that one participant underwent surgery between the end of the intervention and post-examination, giving a dropout rate of 16% in phase A with 95%CI [.016; .304].

### Secondary outcomes

#### Compliance with regard to training recommendations

On average, the participants completed the individual exercise program 3.4 (2.5, 5.6) times per week, ranging from 1.3 to 6.8. The median training time was given as 98 (85, 150) min per week (35 to 304). Endurance training was completed an average of 2.8 (1.9, 4.9) times per week (0 to 6.7), with the median training time per week reaching 167 (86, 247) min (0 to 497 min). Overall, the median weekly training time of the participants was 268 (210, 328) min per week, ranging from 63 to 753 min per week.

#### Adverse events

A total of three participants (12%) reported adverse events that could be related to the exercise. One of these was pain in the Achilles tendon area, one patient complained of knee pain after the first training session and one patient reported pain in the shoulder.

#### Satisfaction with the information about the training

All patients (100%) stated that the information they received about the training was sufficient.

#### Use of the exercise materials

The exercise manual was used by 76% (n = 16) of the participants and the USB stick with the training videos by 62% (n = 13), with 32% (n = 8) stating that they used both. The small equipment (elastic band and inflatable exercise ball) was used by 86% (n = 18) of the participants.

#### Influence of telephone calls on motivation to train

A total of 86% (n = 18) of participants stated that the weekly calls from the therapists had a positive influence (29% very, 24% quite, 33% somewhat) on motivation to train.

## Discussion

To our knowledge, the OSHO #94 study is the first study to investigate the short- and medium-term effects of 100% individualized home training in PwHNC after completion of cancer therapy or in a stable situation under immunotherapy. In contrast to previous studies (28, 43–61), participants do not receive an exercise program with defined FITT criteria (FITT frequency, intensity, time, type) that apply equally to all intervention participants, but rather exercise recommendations based on the current physical activity guidelines for cancer survivors (62). The participants can perform the endurance training according to their preferences, and the recommendations regarding strength and mobility are adapted to the individual’s needs/deficits. OSHO #94 is thus pursuing the approach of transferring the knowledge previously generated primarily in randomized controlled trials (RCT) regarding the effectiveness of targeted exercise interventions in PwHNC (43, 45, 47–60, 63, 64) to ‘real-world’ care. The home-based training approach was chosen (i) because, in our view, it comes closest to the preferences of PwHNC (25–27). (ii) In addition, home-based training programs potentially offer all PwHNC the opportunity to participate, regardless of their place of residence. (iii) In contrast to temporary rehabilitation programs, home-based exercise programs can be continued indefinitely, which can increase QoL and physical activity levels not only in the short but also in the medium term. (iv) The recruiting centres (Rostock, Halle, Greifswald) are located in the northeast of Germany, a rather sparsely populated region with poor infrastructure. Specific exercise programs for people with cancer are scarce in this region, making it difficult to refer PwHNC to community-based programs. (v) Home exercise programs are cost-effective, as there are no membership fees and no travel costs, and they also relieve the burden on the healthcare system/caregiver. (vi) By eliminating the need to travel, home exercise programs are less time consuming and can potentially reduce barriers to exercise (23). Although home exercise programs for PwHNC offer various advantages over group training, feasibility, effectiveness and sustainability have been insufficiently studied, and further research, especially with greater attention to implementation science aspects, seems warranted.

Previous studies with home exercise approaches (65–67) and the feasibility results of OSHO #94 – low dropout and low number of reported adverse events – showed that the chosen home exercise approach is safe to implement. The dropout observed in the feasibility study was clearly below our expectations, which were influenced by the results of Cnossen et al. (68). Among other things, the authors investigated the treatment adherence of PwHNC who participated in a guided home-based prophylactic exercise program during treatment. Adherence was 38% after 12 weeks. The authors saw the main reason for the decrease in adherence in the increasing negative effects of the treatment, which is only to be expected in exceptional cases due to the inclusion criteria in OSHO #94. Since the estimated dropout rate of the OSHO #94 phase A trial is identical to that of our initial study, which we conducted in a group setting, we consider the assumption of 20% dropout to be realistic. Therefore, we adjusted our original calculation of the sample size with a calculated dropout of 30% (N = 60 PwHNC) to 20% (N = 53 PwHNC).

Although, in line with our expectations, the range in terms of training volume (frequency and time) in the feasibility phase is very large, the majority of the PwHNC included are adhering to the exercise recommendations or even exceeding them. One reason for this could lie in the included cohort itself, as the demographic data show that so far mainly PwHNC with a high level of education and a history of sport have been included. The fact that the willingness to participate in an exercise intervention is higher here is consistent with the results of Buffart et al. (69). In addition, it is well known that education/social status and health (behaviour) correlate with each other in the general population (70–72). Another reason could be the study design, which involves weekly telephone calls between therapists and study participants. According to the feasibility results, these telephone calls increase the motivation to exercise in four out of five participants. The results of the FU study will show the influence of the absence of telephone calls on physical activity. Conducting the FU study three months after the end of the intervention will allow an initial assessment of whether the guided home exercise program has an impact on the physical activity behaviour and QoL of PwHNC and can increase the physical activity level in the medium term.

Representatives of the target population were involved in the development of the OSHO #94 study by jointly developing and creating the exercise manual and the training videos. Since the small devices have also been used to a high degree by the current participants, it was decided that the study design would be adhered to, even if the demographic data/lifestyle parameters of the PwHNC included so far do not reflect the typical PwHNC (male, smoker, low level of education). As access to the study was deliberately chosen to be low-threshold – direct approach of the PwHNC or contact via telephone or email from the patients, three on-site appointments over a period of six months with travel costs covered, no costs for materials and support – the results of the study will provide information on which PwHNC are addressed with the approach of guided home exercise.

The selected assessment in the OSHO #94 study corresponds to international standards and includes instruments that were/are regularly used in studies with PwHNC and thus enable comparability of the results. The EORTC QLQ-C30 is used to assess the primary outcome, the QoL, as recommended by Burgos-Mansilla et al. (17). This global instrument appears to be more sensitive to changes in general QoL than to more specific instruments. The GSLTPAQ, an instrument widely used in oncology research, is also used to record and classify physical activity. In accordance with the recommendations of Amireault et al. (36) the original form of the GSLTPAQ is used, and the LSI is used for interpretation.

### Limitations

The design of the OSHO #94 study has some limitations, which are first related to the single-arm design, which does not allow a comparison of the results with a control group (usual care). Second, due to the diverse dissemination of information about the study at three study centres, via regional self-help groups as well as the Germany-wide patient network, social media and flyers, no statement on the recruitment rate is possible. In previous studies with PwHNC, recruitment rates ranged from 20% to 32.3% (17). With regard to recruitment, it should be mentioned that the number of recruiting centres is lower than originally planned due to the COVID-19 pandemic. As a result, recruitment is slower and the recruitment period had to be extended. Third, the LSI of the GSLTPAQ is used to interpret physical activity levels (36). Since PROs for physical activity only correspond to objective measurement data, for example, that collected with accelerometers, to a limited extent (73), this can lead to incorrect classifications of PwHNC. Since the GSLTPAQ is used at all three measurement times, it can be assumed that the ‘errors’ are constant, and thus changes in the activity level can be reliably mapped. The OSHO #94 study will allow us to better understand the effectiveness of guided home exercise programs at the individual level and to evaluate processes to support future implementation and sustainability.

## Declarations

## Data Availability

The data underlying the results presented in the study are available from

## Acknowledgements

Our greatest thanks go to the patients who helped to create the exercise manual and the videos. We would also like to thank the self-help network Kopf-Hals-M.U.N.D.-Krebs e. V. for promoting the study.

## Authors contributions

SF, ÄG, CJ and CGT developed the study concept and protocol. SF, CJ and CGT contributed to the acquisition of funding. SF, JR, LAB, DFS, JS, SS, MB, UK and CGT will oversee the implementation of the protocol and contribute to the acquisition, analysis and interpretation of data. SF, ÄG and CGT drafted the manuscript; JR, LAB and UK contributed to revisions and DFS, JS, SS, MB and CJ approved the final manuscript.

## Data availability statement

The study protocol and the raw data will be made available by the authors without reservation after completion of the study.

## Literature Cited

1. Riechelmann H, Dejaco D, Steinbichler TB, Lettenbichler-Haug A, Anegg M, Ganswindt U et al. Functional Outcomes in Head and Neck Cancer Patients. Cancers (Basel) 2022; 14(9).

2. Lo Nigro C, Denaro N, Merlotti A, Merlano M. Head and neck cancer: improving outcomes with a multidisciplinary approach. Cancer Manag Res 2017; 9:363–71.

3. Ortiz-Comino L, Fernández-Lao C, Speksnijder CM, Lozano-Lozano M, Tovar-Martín I, Arroyo-Morales M et al. Upper body motor function and swallowing impairments and its association in survivors of head and neck cancer: A cross-sectional study. PLoS ONE 2020; 15(6):e0234467.

4. van Hinte G, Leijendekkers RA, Merkx MAW, Takes RP, Nijhuis-van der Sanden MWG, Speksnijder CM. Identifying unmet needs and limitations in physical health in survivors of Head and Neck Cancer. Eur J Cancer Care (Engl) 2021; 30(5):e13434.

5. Baxi SS, Schwitzer E, Jones LW. A review of weight loss and sarcopenia in patients with head and neck cancer treated with chemoradiation. Cancers Head Neck 2016; 1(1):5.

6. Ellis MA, Sterba KR, Brennan EA, Maurer S, Hill EG, Day TA et al. A Systematic Review of Patient-Reported Outcome Measures Assessing Body Image Disturbance in Patients with Head and Neck Cancer. Otolaryngol Head Neck Surg 2019; 160(6):941–54.

7. Couch M, Lai V, Cannon T, Guttridge D, Zanation A, George J et al. Cancer cachexia syndrome in head and neck cancer patients: part I. Diagnosis, impact on quality of life and survival, and treatment. Head Neck 2007; 29(4):401–11.

8. Epstein JB, Robertson M, Emerton S, Phillips N, Stevenson-Moore P. Quality of life and oral function in patients treated with radiation therapy for head and neck cancer. Head Neck 2001; 23(5):389–98.

9. Langendijk JA, Doornaert P, Verdonck-de Leeuw IM, Leemans CR, Aaronson NK, Slotman BJ. Impact of late treatment-related toxicity on quality of life among patients with head and neck cancer treated with radiotherapy. Journal of clinical oncology 2008; 26(22):3770–6.

10. Chow LQM. Head and Neck Cancer. The new england journal of medicine 2020; 382(1):60–72.

11. León X, Orús C, Casasayas M, Neumann E, Holgado A, Quer M. Trends in disease-specific survival of head and neck squamous cell carcinoma patients treated in a single institution over a 30-year period. Oral Oncol 2021; 115:105184.

12. Ferris RL, Blumenschein G, Fayette J, Guigay J, Colevas AD, Licitra L et al. Nivolumab for Recurrent Squamous-Cell Carcinoma of the Head and Neck. N Engl J Med 2016; 375(19):1856–67.

13. Ringash J, Bernstein LJ, Devins G, Dunphy C, Giuliani M, Martino R et al. Head and Neck Cancer Survivorship: Learning the Needs, Meeting the Needs. Semin Radiat Oncol 2018; 28(1):64–74.

14. Margalit DN, Salz T, Venchiarutti R, Milley K, McNamara M, Chima S et al. Interventions for head and neck cancer survivors: Systematic review. Head Neck 2022; 44(11):2579–99.

15. Burgos-Mansilla B, Galiano-Castillo N, Lozano-Lozano M, Fernández-Lao C, Lopez-Garzon M, Arroyo-Morales M. Effect of Physical Therapy Modalities on Quality of Life of Head and Neck Cancer Survivors: A Systematic Review with Meta-Analysis. J Clin Med 2021; 10(20).

16. Pérez IMM, Pérez SEM, García RP, Lupgens DdZ, Martínez GB, González CR et al. Exercise-based rehabilitation on functionality and quality of life in head and neck cancer survivors. A systematic review and meta-analysis. Sci Rep 2023; 13(1):8523.

17. Avancini A, Borsati A, Belluomini L, Giannarelli D, Nocini R, Insolda J et al. Effect of exercise across the head and neck cancer continuum: a systematic review of randomized controlled trials. Support Care Cancer 2023; 31(12):670.

18. Bye A, Sandmael JA, Stene GB, Thorsen L, Balstad TR, Solheim TS et al. Exercise and Nutrition Interventions in Patients with Head and Neck Cancer during Curative Treatment: A Systematic Review and Meta-Analysis. Nutrients 2020; 12(11).

19. Capozzi LC, Nishimura KC, McNeely ML, Lau H, Culos-Reed SN. The impact of physical activity on health-related fitness and quality of life for patients with head and neck cancer: a systematic review. Br J Sports Med 2016; 50(6):325–38.

20. Goyal N, Day A, Epstein J, Goodman J, Graboyes E, Jalisi S et al. Head and neck cancer survivorship consensus statement from the American Head and Neck Society. Laryngoscope Investig Otolaryngol 2022; 7(1):70–92.

21. Fang Y-Y, Wang C-P, Chen Y-J, Lou P-J, Ko J-Y, Lin J-J et al. Physical activity and fitness in survivors of head and neck cancer. Support Care Cancer 2021; 29(11):6807–17.

22. Karczewska-Lindinger M, Tuomi L, Fridolfsson J, Arvidsson D, Börjesson M, Finizia C. Low physical activity in patients diagnosed with head and neck cancer. Laryngoscope Investig Otolaryngol 2021; 6(4):747–55.

23. Ning Y, Wang Q, Ding Y, Zhao W, Jia Z, Wang B. Barriers and facilitators to physical activity participation in patients with head and neck cancer: a scoping review. Support Care Cancer 2022; 30(6):4591–601.

24. Doughty HC, Hill RA, Riley A, Midgley AW, Patterson JM, Boddy LM et al. Barriers to and facilitators of physical activity in adults living with and beyond cancer, with special emphasis on head and neck cancer: a systematic review of qualitative and mixed methods studies. Support Care Cancer 2023; 31(8):471.

25. Midgley AW, Lowe D, Levy AR, Mepani V, Rogers SN. Exercise program design considerations for head and neck cancer survivors. European archives of oto-rhino-laryngology 2018; 275(1):169–79.

26. Rogers LQ, Malone J, Rao K, Courneya KS, Fogleman A, Tippey A et al. Exercise preferences among patients with head and neck cancer: prevalence and associations with quality of life, symptom severity, depression, and rural residence. Head Neck 2009; 31(8):994–1005.

27. Jackson C, Dowd AJ, Capozzi LC, Bridel W, Lau HY, Culos-Reed SN. A turning point: Head and neck cancer patients’ exercise preferences and barriers before and after participation in an exercise intervention. Eur J Cancer Care (Engl) 2018; 27(2):e12826.

28. Felser S, Behrens M, Strüder D, Liese J, Rohde K, Junghanss C et al. Feasibility and Effects of a Supervised Exercise Program Suitable for Independent Training at Home on Physical Function and Quality of Life in Head and Neck Cancer Patients: A Pilot Study. Integr Cancer Ther 2020; 19:1–12. Available from: URL:.

29. Felser S, editor. Übungshandbuch für Patienten mit Mund-, Kiefer-, Gesichts- und Halstumoren. Rostock; 2019.

30. Borg GA. Psychophysical bases of perceived exertion. Med Sci Sports Exerc 1982; 14(5):377–81.

31. Bjordal K, Graeff A de, Fayers PM, Hammerlid E, van Pottelsberghe C, Curran D, et al. A 12 country field study of the EORTC QLQ-C30 (version 3.0) and the head and neck cancer specific module (EORTC QLQ-H&N35) in head and neck patients. European Journal of Cancer 2000; 36:1796–1807.

32. Aaronson NK, Ahmedzai S, Bergman B, Bullinger M, Cull A, Duez NJ et al. The European Organization for Research and Treatment of Cancer QLQ-C30: a quality-of-life instrument for use in international clinical trials in oncology. J Natl Cancer Inst 1993; 85(5):365–76.

33. Fayers PM. Interpreting quality of life data: population-based reference data for the EORTC QLQ-C30. European Journal of Cancer 2001; 37(11):1331–4.

34. Singer S, Araújo C, Arraras JI, Baumann I, Boehm A, Brokstad Herlofson B et al. Measuring quality of life in patients with head and neck cancer: Update of the EORTC QLQ-H&N Module, Phase III. Head Neck 2015; 37(9):1358–67.

35. Amireault S, Godin G. The Godin-Shephard leisure-time physical activity questionnaire: validity evidence supporting its use for classifying healthy adults into active and insufficiently active categories. Percept Mot Skills 2015; 120(2):604–22.

36. Amireault S, Godin G, Lacombe J, Sabiston CM. The use of the Godin-Shephard Leisure-Time Physical Activity Questionnaire in oncology research: a systematic review. BMC Med Res Methodol 2015; 15:60.

37. Godin G. The Godin-Shephard Leisure-Time Physical Activity Questionnaire. The Health & Fitness Journal of Canada 2011; 4(1):18–22.

38. Mâsse LC, Niet JE de. Sources of validity evidence needed with self-report measures of physical activity. J Phys Act Health 2012; 9 Suppl 1:S44–55.

39. Holt LE, Pelham TW, Burke DG. Modifications to the Standard Sit-and-Reach Flexibility Protocol. J Athl Train 1999; 34(1):43–7.

40. Guralnik JM, Ferrucci L, Pieper CF, Leveille SG, Markides KS, Ostir GV et al. Lower extremity function and subsequent disability: consistency across studies, predictive models, and value of gait speed alone compared with the short physical performance battery. J Gerontol A Biol Sci Med Sci 2000; 55(4):M221–31.

41. Schmidt K, Vogt L, Thiel C, Jäger E, Banzer W. Validity of the six-minute walk test in cancer patients. Int J Sports Med 2013; 34(7):631–6.

42. Borg E, Borg G, Larsson K, Letzter M, Sundblad B-M. An index for breathlessness and leg fatigue. Scand J Med Sci Sports 2010; 20(4):644–50.

43. Bragante KC, Groisman S, Carboni C, Baiocchi JMT, da Motta NW, Silva MF et al. Efficacy of exercise therapy during radiotherapy to prevent reduction in mouth opening in patients with head and neck cancer: A randomized controlled trial. Oral Surg Oral Med Oral Pathol Oral Radiol 2020; 129(1):27–38.

44. Capozzi LC, Boldt KR, Lau H, Shirt L, Bultz B, Culos-Reed SN. A clinic-supported group exercise program for head and neck cancer survivors: managing cancer and treatment side effects to improve quality of life. Sports medicine 2015; 23(4):1001–7.

45. Capozzi LC, McNeely ML, Lau HY, Reimer RA, Giese-Davis J, Fung TS et al. Patient-reported outcomes, body composition, and nutrition status in patients with head and neck cancer: Results from an exploratory randomized controlled exercise trial. Cancer 2016; 122(8):1185–200.

46. Crevenna R, Schneider B, Mittermaier C, Keilani M, Zöch C, Nuhr M et al. Implementation of the Vienna Hydrotherapy Group for Laryngectomees--a pilot study. Support Care Cancer 2003; 11(11):735–8.

47. Dotevall H, Tuomi L, Petersson K, Löfhede H, Bergquist H, Finizia C. Treatment with head-lift exercise in head and neck cancer patients with dysphagia: results from a randomized, controlled trial with flexible endoscopic evaluation of swallowing (FEES). Support Care Cancer 2022; 31(1):56.

48. Grote M, Maihöfer C, Weigl M, Davies-Knorr P, Belka C. Progressive resistance training in cachectic head and neck cancer patients undergoing radiotherapy: a randomized controlled pilot feasibility trial. Radiat Oncol 2018; 13(1):215.

49. Hajdú SF, Wessel I, Dalton SO, Eskildsen SJ, Johansen C. Swallowing Exercise During Head and Neck Cancer Treatment: Results of a Randomized Trial. Dysphagia 2022; 37(4):749–62.

50. Lin K-Y, Cheng H-C, Yen C-J, Hung C-H, Huang Y-T, Yang H-L et al. Effects of Exercise in Patients Undergoing Chemotherapy for Head and Neck Cancer: A Pilot Randomized Controlled Trial. Int J Environ Res Public Health 2021; 18(3).

51. Loh E-W, Shih H-F, Lin C-K, Huang T-W. Effect of progressive muscle relaxation on postoperative pain, fatigue, and vital signs in patients with head and neck cancers: A randomized controlled trial. Patient Educ Couns 2022; 105(7):2151–7.

52. Lønbro S, Dalgas U, Primdahl H, Johansen J, Nielsen JL, Aagaard P et al. Progressive resistance training rebuilds lean body mass in head and neck cancer patients after radiotherapy--results from the randomized DAHANCA 25B trial. Radiother Oncol 2013; 108(2):314–9.

53. Lønbro S, Dalgas U, Primdahl H, Overgaard J, Overgaard K. Feasibility and efficacy of progressive resistance training and dietary supplements in radiotherapy treated head and neck cancer patients-the DAHANCA 25A study. Acta oncologica 2013; 52(2):310–8.

54. McGarvey AC, Hoffman GR, Osmotherly PG, Chiarelli PE. Maximizing shoulder function after accessory nerve injury and neck dissection surgery: A multicenter randomized controlled trial. Head Neck 2015; 37(7):1022–31.

55. McNeely ML, Parliament M, Courneya KS, Seikaly H, Jha N, Scrimger R et al. A pilot study of a randomized controlled trial to evaluate the effects of progressive resistance exercise training on shoulder dysfunction caused by spinal accessory neurapraxia/neurectomy in head and neck cancer survivors. Head Neck 2004; 26(6):518–30.

56. McNeely ML, Parliament MB, Seikaly H, Jha N, Magee DJ, Haykowsky MJ et al. Effect of exercise on upper extremity pain and dysfunction in head and neck cancer survivors: a randomized controlled trial. Cancer 2008; 113(1):214–22.

57. Rogers LQ, Anton PM, Fogleman A, Hopkins-Price P, Verhulst S, Rao K et al. Pilot, randomized trial of resistance exercise during radiation therapy for head and neck cancer. Head Neck 2013; 35(8):1178–88.

58. Samuel SR, Maiya GA, Babu AS, Vidyasagar MS. Effect of exercise training on functional capacity & quality of life in head & neck cancer patients receiving chemoradiotherapy. Indian J Med Res 2013; 137(3):515–20.

59. Thomas A, D’Silva C, Mohandas L, Pais SMJ, Samuel SR. Effect of Muscle Energy Techniques V/S Active Range of Motion Exercises on Shoulder Function Post Modified Radical Neck Dissection in patients with Head and Neck Cancer - A Randomized Clinical Trial. Asian Pac J Cancer Prev 2020; 21(8):2389–93.

60. Valkenet K, Trappenburg JCA, Ruurda JP, Guinan EM, Reynolds JV, Nafteux P et al. Multicentre randomized clinical trial of inspiratory muscle training versus usual care before surgery for oesophageal cancer. Br J Surg 2018; 105(5):502–11.

61. Zhao SG, Alexander NB, Djuric Z, Zhou J, Tao Y, Schipper M et al. Maintaining physical activity during head and neck cancer treatment: Results of a pilot controlled trial. Head Neck 2016; 38 Suppl 1:E1086–96.

62. Campbell KL, Winters-Stone KM, Wiskemann J, May AM, Schwartz AL, Courneya KS et al. Exercise Guidelines for Cancer Survivors: Consensus Statement from International Multidisciplinary Roundtable. Med Sci Sports Exerc 2019; 51(11):2375–90.

63. Su T-L, Chen A-N, Leong C-P, Huang Y-C, Chiang C-W, Chen I-H et al. The effect of home-based program and outpatient physical therapy in patients with head and neck cancer: A randomized, controlled trial. Oral Oncol 2017; 74:130–4.

64. Hong Y-L, Hsieh T-C, Chen P-R, Chang S-C. Nurse-Led Counseling Intervention of Postoperative Home-Based Exercise Training Improves Shoulder Pain, Shoulder Disability, and Quality of Life in Newly Diagnosed Head and Neck Cancer Patients. J Clin Med 2022; 11(14).

65. Kok A, Passchier E, May AM, van den Brekel MWM, Jager-Wittenaar H, Veenhof C, et al. Feasibility of a supervised and home-based tailored exercise intervention in head and neck cancer patients during chemoradiotherapy. Eur J Cancer Care (Engl) 2022; 31(6):e13662.

66. Cnossen IC, van Uden-Kraan CF, Rinkel RNPM, Aalders IJ, Goede CJT de, Bree R de, et al. Multimodal guided self-help exercise program to prevent speech, swallowing, and shoulder problems among head and neck cancer patients: a feasibility study. J Med Internet Res 2014; 16(3):e74.

67. Eades M, Murphy J, Carney S, Amdouni S, Lemoignan J, Jelowicki M et al. Effect of an interdisciplinary rehabilitation program on quality of life in patients with head and neck cancer: review of clinical experience. Head Neck 2013; 35(3):343–9.

68. Cnossen IC, van Uden-Kraan CF, Witte BI, Aalders YJ, Goede CJT de, Bree R de, et al. Prophylactic exercises among head and neck cancer patients during and after swallowing sparing intensity modulated radiation: adherence and exercise performance levels of a 12-week guided home-based program. Eur Arch Otorhinolaryngol 2017; 274(2):1129–38.

69. Buffart LM, Bree R de, Altena M, van der Werff S, Drossaert CHC, Speksnijder CM, et al. Demographic, clinical, lifestyle-related, and social-cognitive correlates of physical activity in head and neck cancer survivors. Support Care Cancer 2018; 26(5):1447–56.

70. Hannover B, Kleiber D. Gesundheit und Bildung. In: Tippelt R, Schmidt-Hertha B, editors. Handbuch Bildungsforschung. Wiesbaden: Springer Fachmedien Wiesbaden; 2016. p. 1–16.

71. Raghupathi V, Raghupathi W. The influence of education on health: an empirical assessment of OECD countries for the period 1995–2015. Archives of public health 2020; 78(1):1–18.

72. Zajacova A, Lawrence EM. The Relationship Between Education and Health: Reducing Disparities Through a Contextual Approach. Annu Rev Public Health 2018; 39:273–89.

73. Douma JAJ, Beaufort MB de, Kampshoff CS, Persoon S, Vermaire JA, Chinapaw MJ, et al. Physical activity in patients with cancer: self-report versus accelerometer assessments. Support Care Cancer 2020; 28(8):3701–9.

